# The cross-sectional study of hospitalized coronavirus disease 2019 patients in Xiangyang, Hubei province

**DOI:** 10.1101/2020.02.19.20025023

**Authors:** Jin-Wei Ai, Jun-Wen Chen, Yong Wang, Xiao-Yun Liu, Wu-Feng Fan, Gao-Jing Qu, Mei-Ling Zhang, Sheng-Duo Pei, Bo-Wen Tang, Shuai Yuan, Yang Li, Li-Sha Wang, Guo-Xin Huang, Bin Pei

## Abstract

**Objective:** To describe the epidemiological and clinical characteristics of the Coronavirus Disease 2019 (COVID-19) hospitalized patients and to offer suggestions to the urgent needs of COVID-19 prevention, diagnosis and treatment.

**Methods:** We included 102 confirmed COVID-19 cases hospitalized in Xiangyang No.1 people’s hospital, Hubei, China until Feb 9th, 2020. Demographic data, laboratory findings and chest computed tomographic (CT) images were obtained and analyzed.

**Findings:** All cases were confirmed by real-time RT-PCR, including 52 males and 50 females with a mean age of 50.38 years (SD 16.86). Incubation time ranged from one to twenty days with a mean period of 8.09 days (SD 4.99). Fever (86[84.3%] of 102 patients), cough (58[57%]), fatigue (28[27%]), shortness of breath (24[23%]), diarrhea (15[15%]), expectoration (13[12%]), inappetence (11[10%]) were common clinical manifestations. We observed a decreased blood leukocyte count and lymphopenia in 21 (20.6%) and 56 (54.9%) patients, respectively. There were 66 (68%) of 97 patients with elevated C-reactive protein levels and 49 (57.6%) of 85 with increased erythrocytes sedimentation rate. Higher levels of procalcitonin and ferritin were observed in 19 (25.3%) of 75 and 12 (92.3%) of 13 patients, respectively. Eight patients were admitted to intensive care unit (ICU), six developed respiratory failure, three had multiple organ failure and three died. The cumulative positivity rate over three rounds of real-time RT-PCR was 96%. One-hundred patients were found with typical radiological abnormalities in two rounds of chest CT scans, indicating a 98% consistency with real-time RT-PCR results.

**Interpretation:** Most COVID-19 patients in Xiangyang were secondary cases without sex difference, and the rate of severe case and death was low. Middle-to-old-age individuals were more susceptible to the virus infection and the subsequent development of severe/fatal consequences. The average incubation period was longer among our patients. We recommend prolonging the quarantine period to three weeks. Three times real-time RT-PCR plus two times CT scans is a practical clinical diagnosis strategy at present and should be used to increase the accuracy of diagnosis, thereby controlling the source of infection more effectively.

## Introduction

In December 2019, a novel coronavirus-infected pneumonia epidemic outbroke in Wuhan, Hubei province, China. It has spread to all over China and many other countries within a short period of time^[1, 2]^. The virus was later whole-genome sequenced and identified as a new coronavirus, named severe acute respiratory syndrome coronavirus 2 (SARS-CoV-2). The syndrome caused by SARS-CoV-2 infection was called as COVID-19, short for “coronavirus disease 2019” by World Health Organization^[3]^.

Coronaviruses (CoVs) are the viruses with an enveloped positive-sense single-stranded ribonucleic acid (RNA) genome^[4]^. The name comes from the typical appearance of the virions under the electron microscopy that they all have glycoprotein spike on the surface which resembles the shape of crown. CoVs exist extensively in the nature, could infect birds and mammals, including human^[5, 6]^. CoVs are known to spread through close contact, especially through respiratory droplets during cough and sneeze. Most infections only cause mild respiratory symptoms like common cold. Severe cases could develop acute respiratory distress syndrome, multiple organs failure, and even fatalities^[7]^. CoVs could be divided into α, β, γ and δ genera. SARS-CoV-2 belongs to the β-coronavirus together with the severe acute respiratory syndrome coronavirus (SARS-CoV) that outbroke in 2003 and the Middle East respiratory syndrome coronavirus (MERS-CoV) that outbroke in 2012^[8]^.

Until February 11, 2020, a total number of 38 800 COVID-19 cases have been diagnosed in China, among which 8 204 were severe cases and 1 113 deaths have been reported. COVID-19 has become a great threat to human health and cause great burden to the whole society. Further exploration of SRAS-CoV-2 would benefit COVID-19 prevention, diagnosis and treatment. Although Huang *et al*^[9]^ and Chen *et al*^[10]^ uncovered some basic epidemiological and clinical features of COVID-19 patients in Wuhan city, data for the patients from other regions in Hubei Province were still scarce. Xiangyang is the second biggest city in Hubei with a large number of labor-force migrant to Wuhan. At the moment, Xiangyang had many diagnosed cases already. In order to understand the features of COVID-19 in Xiangyang, we conducted a cross-sectional study to describe the epidemiological and clinical characteristics of COVID-19 patients hospitalized at Xiangyang No.1 people’s hospital.

## Methods

### Study design and participants

We recruited patients with positive real-time RT-PCR results who were admitted to Xiangyang No.1 People’s Hospital before February 9th, 2020. Data were collected until February 10th, 2020. Diagnosis was made by three physicians based on *Diagnosis Guidance for Novel Coronavirus Pneumonia the 5*^*th*^ *Edition* published by National Health Commission of the People’s Republic of China and National Administration of Traditional Chinese Medicine^[11]^. The current study was approved by the ethics review board at Xiangyang No.1 People’s Hospital (No. 2020GCP012).

### Data collection

Data were extracted by two groups (three physicians per group) from the hospital information system using a consistent data collection protocol and cross-checked. We collected following information: demographic information, exposure history, medical history, principal clinical symptoms and their onset time, real-time RT-PCR results, laboratory findings, radiological findings, comorbidities and disease progression. Laboratory examination included blood routine and blood biochemistry such as alanine transaminase (ALT), aspartate aminotransferase (AST), creatine kinase (CK), creatine kinase isoenzymes-MB (CK-MB), lactate dehydrogenase (LDH), α-hydroxybutyrate dehydrogenase (α-HBDH), C-reactive protein (CRP), procalcitonin (PCT), erythrocyte sedimentation rate (ESR) and serum ferritin.

Positive real-time RT-PCR was defined as having at least one time of positive result at our hospital or other hospitals. Diagnostic results of chest computed tomographic (CT) scans was given by two radiologists independently, and then were cross-checked. For the patients with inconsistent diagnosis results or were suspected, final diagnosis was made after the deliberation of two radiologists. Only first laboratory findings and CT scan results after hospitalization were used in the present study.

### Statistical analysis

Categorical data were described using frequency and percentage. Normalibility of continuous data was tested. We used mean (standard deviation, SD) to describe variables with normal distribution, otherwise median were used. All the analyses were performed in Stata 14.

## Results

### Epidemiological characteristics

A total of 102 cases (52 males and 50 females) with positive real-time RT-PCR results were included in our study. The mean age was 50.38 (16.86) years old, ranging from 1.5 to 90. The majority of cases fell into the age group of 50-70 years old. Among 71 cases with confirmed contact history, seven were Wuhan residents, 37 had travel history to Wuhan, four contacted with diagnosed patients, and 23 were family-clustered cases. In the analysis of 44 cases with clear contact time, incubation periods ranged from one to twenty days with a mean of 8.09 (4.99) days (Table 1).

**Table 1.** Baseline characteristics and clinical outcomes of patients with COVID-19.

|  |  | Patients (n=102) |
| --- | --- | --- |
| Sex | Male | 52 (51%) |
|  | Female | 50 (49%) |
| Age, years | Mean (SD) | 50.38 (16.86) |
|  | Range | 1.5-90 |
|  | < 18 | 2 (2%) |
|  | 18-30 | 9 (8.8%) |
|  | 30-40 | 16 (15.7%) |
|  | 40-50 | 21 (20.6%) |
|  | 50-60 | 23 (22.5%) |
|  | 60-70 | 16 (15.7%) |
|  | 70-80 | 12 (11.8%) |
|  | ≥80 | 3 (2.9%) |
| Exposure history | Imported | 7 (6.9%) |
|  | Travel history to Wuhan | 37 (36.3%) |
|  | Contacts with patients | 4 (3.9%) |
|  | Family clustering | 23 (22.5%) |
|  | Unknown | 31 (30.4%) |
| Chronic illness |  | 39 (38.2%) |
| Signs and symptoms | Fever | 86 (84.3%) |
|  | Cough | 58 (56.9%) |
|  | Sputum production | 13 (12.7%) |
|  | Chill | 12 (11.8%) |
|  | Myalgia | 3 (2.9%) |
|  | Fatigue | 28 (27.5%) |
|  | Palpitations | 2 (2.0%) |
|  | Stomachache | 3 (2.9%) |
|  | Diarrhea | 15 (14.3%) |
|  | Nausea | 9 (8.8%) |
|  | Vomit | 2 (2.0%) |
|  | Inappetence | 11 (10.8%) |
|  | Sore throat | 6 (5.9%) |
|  | Chest pain | 3 (2.9%) |
|  | Dizziness | 4 (3.9%) |
|  | Shortness of breath | 24 (23.5%) |
|  | Respiratory failure | 9 (8.8%) |
| Clinical outcome | Death | 3(3%) |
|  | Cure | 7(7%) |

Data are n (%) unless specified otherwise. COVID-19 = coronavirus disease 2019.

Thirty-nine cases (38.2%) had comorbidities, including hypertension, diabetes, coronary artery disease, tuberculosis, hepatitis B and chronic bronchitis. Eight cases (7.9%) were admitted to intensive care unit (ICU). Among those, six cases acquired respiratory failure, three developed organ failure and three died. The fatality rate was 3%. After two weeks of treatment, symptoms of seven (7%) patients subsided and CT image showed clear absorption of ground-glass opacity in lungs, negativities detected in two rounds of virus RNA detections. However, they are still being quarantined in case of false negativity.

### Real -time RT-PCR

All suspected cases were performed with real-time RT-PCR. The next test was performed in other day in cases with negative result. There are 67 (66%) cases with positive results in the first round of test, 21 were found in the second-round with cumulative positivity rate of 86%, 10 were found in the third-round with cumulative positivity rate of 96%, and 2 cases were in the fourth and fifth round, respectively.

### Chest CT scan

In 102 cases confirmed by real-time RT-PCR, 90 cases had typical radiological abnormalities in first chest CT scan, which indicated an 88% consistency with molecular test. In the 11 cases who took re-examination after 5-7days, 10 cases showed radiological abnormalities. Taking together, CT scan had a 98% diagnostic rate consistent with real-time RT-PCR.

The mild radiographic abnormalities demonstrated as sparse subpleural nodular or patchy ground-glass opacities that principally distributed in lung segments and under pleura, with clear borderline and vascular shadows showing around (Figure 1 A). Moderate abnormalities were characterized with increased number of ground-glass opacities and their expansion to multiple lobes. Nodular ground-glass opacities occurred with consolidation, with ambiguous borderline and blood vessels going through (Figure 1 B-C). In severe abnormalities, diffuse patchy or grid ground-glass opacities with heterogeneous density presented, partially with clear borderline, in which air bronchogram and vascular shadow can be seen. Both lungs showed white lung change, with thickening of interlobular septum and multiple consolidations in mediastinal window (Figure 1 D).

**Figure 1.**
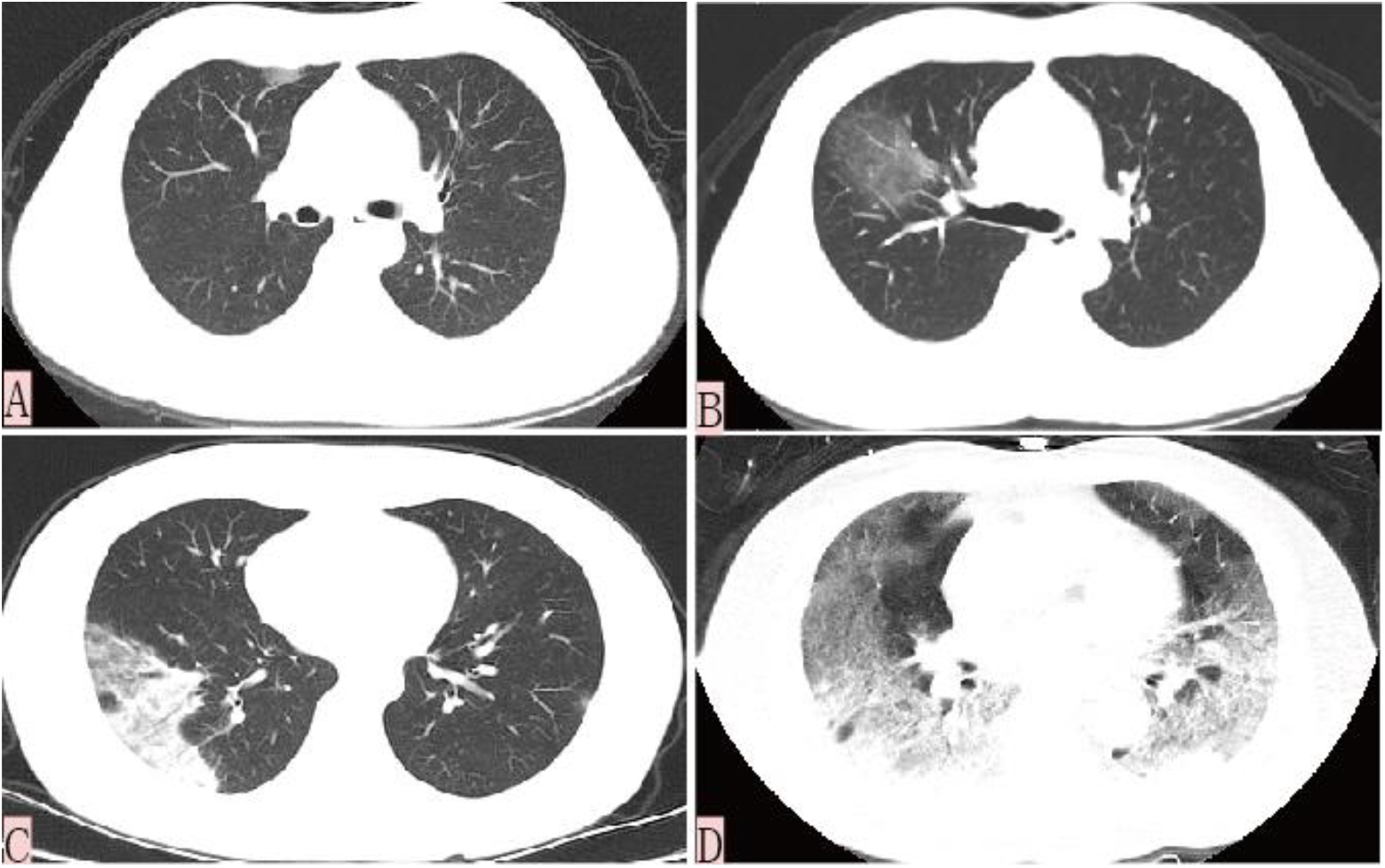
Chest computed tomographic images of patients with coronavirus disease 2019. (A) Mild pneumonia patient showed patchy ground-glass opacity with clear borderline in a transverse chest CT image. (B-C) Transverse chest CT images of moderate patients showing large ground-glass opacities and some with ambiguous borderline. (D) Severe cases were characterized by typical white lung change as high-density mass shadows and multiple lobular consolidations were observed.

### Principal clinical symptoms

Of 102 patients with COVID-19, 86 (84.3%) cases experienced fever, ranging from 37.2 °C to 38.5 °C, with only two cases had extreme body temperature over 39 °C. Fifty-eight (56.9%) cases had cough, mainly dry cough. Ten (9.8%) had white sputum and two (2%) had purulent sputum. Chest X-ray, clinical signs and laboratory examinations indicated that purulent sputum was caused by co-infection with bacteria in lung. Twenty-eight (27.5%) cases showed fatigue and myalgia. Twenty-four (23.5%) cases had respiratory problem, mainly shown as chest tightness, asthma and shortness of breath. Nine (8.8%) cases developed respiratory failure. Fifteen (14.3%) cases showed diarrhea and seven (7%) cases had decreased appetite (Table 1). Two (2%) cases had diarrhea as first appeared symptom, while the other had fever and/or respiratory symptoms as the first manifestation.

### Laboratory findings

Some cases had increased ALT, AST, CK, CK-MB, LDH, α-HBDH. Nineteen (25.3%) cases had increased PCT of 75 patients tested; Thirteen cases underwent ferritin test of which 12 (92.3%) increased (Table 2). Of all 102 patients, leukocyte count decreased in 21 (21.6%) cases and lymphocyte count decreased in 56 (54.9%) cases; CRP increased in 66 (68%) cases of 97 patients; ESR became higher in 49 (57.6%) cases among 85 patients (Table 3).

**Table 2.** Blood biochemistry of 102 patients with COVID-19.

|  | Normal range | Increased | Mean (SD) |
| --- | --- | --- | --- |
| Alanine transaminase, IU/L | 9-50 | 20 (19.6%) | 27.77 (21.13) |
| Aspartate aminotransferase, IU/L | 15-40 | 26 (25.5%) | 30.59 (15.03) |
| Creatine kinase, U/L | 40-200 | 19 (18.6%) | 139.32 (121.66) |
| Creatine kinase-MB, U/L | 0-24 | 11 (10.8%) | 13.94 (10.25) |
| Lactate dehydrogenase, U/L | 120-250 | 37 (36.3%) | 245.38 (14.35) |
| α-Hydroxybutyrate dehydrogenase, U/L | 72-182 | 37 (36.3%) | 178.85 (70.93) |
| Myoglobin, ng/mL* | 0-80 | 4/59 (6.8%) | 39.61 (30.87) |
| Serum ferritin, ng/mL* | 27-375 | 12/13 (92.3%) | 269.89 (124.88) |

Data are n (%) unless specified otherwise. COVID-19 = coronavirus disease 2019. \*Data available as indicated.

**Table 3.** Blood routine and infection-related biomarkers of 102 patients with COVID-19.

|  | Normal range | Increased | Decreased | Mean (SD) |
| --- | --- | --- | --- | --- |
| Absolute leucocytes, $\times 10^9/L$ | 3.5-9.5 | 2 (2%) | 21 (20.6%) | 4.84 (1.62) |
| Neutrophil, % | 40-75 | 22 (21.6%) | 2 (2%) | 65.00 (12.44) |
| Absolute neutrophil, $\times 10^9/L$ | 1.8-6.3 | 4 (3.9%) | 13 (12.7%) | 3.21 (1.43) |
| Lymphocyte, % | 20-50 | 1 (1%) | 36 (35.3%) | 25.24 (11.16) |
| Absolute lymphocyte, $\times 10^9/L$ | 1.1-3.2 | 1 (1%) | 56 (54.9%) | 1.18 (0.73) |
| Monocyte, % | 3-10 | 27 (26.5%) | 0 (0%) | 8.98 (3.40) |
| Absolute monocyte, $\times 10^9/L$ | 0.1-0.6 | 14 (13.7%) | 1 (1%) | 0.41 (0.19) |
| C-reactive protein, mg/L * | 0-8 | 66/97 (68%) | — | 28.16 (26.72) |
| Procalcitonin, ng/mL * | <0.1 | 19/75 (25.3%) | — | — |
| Erythrocyte sedimentation rate, mm/h * | 0-20 | 49/85 (57.6%) | — | 33.30 (23.39) |

Data are n (%) unless specified otherwise. COVID-19 = coronavirus disease 2019. \*Data available as indicated.

### Other pathogens screening

All cases underwent common etiological examination, including MR-IgM, CP-IgM, TBAb-IgG, ADV-IgM, RSV-RNA, H7, Flu A and Flu B. Results were negative in most patients. Only two cases were detected to be weak positive and positive in MR-IgM, respectively. Three cases showed weakly positive and one showed positive in CP-IgM. Additionally, three cases without tuberculosis history were tested to be positive in TBAb-IgG.

## Discussion

SARS-CoV-2 is a highly infective virus. Although strict control measures were taken in Wuhan, Hubei, there were still a large number of new cases every day. Further understanding of the epidemiological and clinical features of COVID-19 is of great importance for the early detection of patients and reduction of severe cases and deaths^[12]^.

The present study was based on 102 diagnosed SARS-CoV-2 infected patients in Xiangyang No.1 People’s Hospital, Hubei. There was no significant sex difference among our cases with a men-to-women ratio of 1.04 (52:50), which is not in line with previous studies. Huang *et al*^[9]^ and Chen *et al*^[10]^ found a higher percentage of men with a men-to-women ratio of 2.71 (30:11) and 2.09 (67:32) in a total of 41 and 99 inpatients, respectively. Discrepancy may be attributed by the differences of the occupational feature between the first- and second-generation transmission cases. In Huang *et al* ^[9]^ study, 27 out of 41 patients were directly exposed to the seafood market in Wuhan where was suspected to be the source of infection. However, in Xiangyang area, the patients were mostly secondary cases. In the process of transmission, the sex difference was gradually diluted, indicating that there might be no gender difference in being infected.

Seven cases were imported from Wuhan and 37 had travel history to Wuhan, together accounting for 43% of 102 cases. The rest were secondary transmission cases, among which 23 were family clustering and 31 patients without clear contact history. Compared with previous studies^[9, 10]^, the main routes of transmission in our cases were different. The family is the fundamental unit of society in China. It is unlikely to avoid close contact without clear diagnosis of the disease, or even if the diagnosis is clear but the patient is not isolated. Based on the family-clustering feature of SARS-CoV-2 infected population in Xiangyang, it is of great significance to confirm suspected cases and quarantine them from their family as early as possible, especially in areas dominated by secondary transmission cases^[13, 14]^.

In our analysis of 44 patients with clear contact history, we found that the mean incubation period of COVID-19 was 8.08 (5.06) days and ranged from 1 to 20 days. Compared with previous studies^[9, 10]^, the incubation period of our patients varied more greatly with maximum of 20 days. The prolonged incubation period will increase the risk of virus transmission. According to *Diagnosis Guidance for Novel Coronavirus Pneumonia the 5*^*th*^ *Edition*^[11]^, individuals with close contact history need to be quarantined for 14 days. The present study suggested that those potential patients were still likely to transmit the virus even after a 14-day quarantine. Real-time RT-RNA testing and medical observation before release should be strengthened and the quarantine time should be extended if necessary.

Only two cases were under 18 years old (one case with 15 and 1.5 years old), which indicated that people with good health status and immunity were not susceptible. The majority of our patients were 50 to 70 years old with common chronic diseases, such as hypertension, diabetes, tuberculosis, hepatitis B, chronic bronchitis, etc. Those people have low immunity, organic damage, and decreased body regulation, which may explain the high incidence of SARS-CoV-2 infection in this age group^[15]^. In addition, disease severity and fatality were higher among the elderly compared with the rest cases. The three deaths in this study were 72, 73 and 78 years old. Among three deaths, one was infected with SARS-CoV-2 after recent lung cancer surgery, one had diabetes, ischemic cardiomyopathy, and hyperthyroidism, and one had hypertension and coronary heart disease^[16, 17]^. In the absence of cure, it is essential to protect the elderly population, and therefore, to reduce the incidence. Meanwhile, early intervention and control of basic diseases may be a way to reduce the critical ill rate and mortality of the aged-infected population.

SARS-CoV-2 mainly causes lower respiratory tract infections^[18]^. Among 102 patients, the major clinical characteristics were fever (84.3%), cough (56.9%), fatigue (27.5%), etc. Most cases had mild fever (body temperature 37.3 °C −38.5 °C) and irritant dry cough. A few patients (12.7%) have a small amount of white frothy sputum and runny nose (2%). In our patients, two cases had diarrhea as the main symptom and 13 cases had diarrhea during the course of the disease. The total incidence of diarrhea and loss of appetite was 15% and 10%, respectively, which differed from previous studies^[9, 10]^. The disagreement may be caused by the nature of the systemic infection of the virus, the insufficient understanding of the disease. A recent study reported that ACE-2-expressingcolonocytes are vulnerable for SARS-CoV-2 infections^[19]^.

The positive rate of the first throat swabs real-time RT-RNA test in patients was 66%; the cumulative positivity rate in two rounds was 86%, and the cumulative positivity rate in three rounds was 96%. Therefore, the sensitivity of virus RNA test as the main diagnostic method is not high, could be due to the low viral load in upper respiratory tract^[20, 21]^. In the absence of more sensitive diagnostic methods, we recommend ≥3 repeated molecular tests to increase the diagnostic rate, in order to effectively control transmission.

Accumulative positivity rate of twice chest CT was 98% consistent with real-time RT-PCR, which means the vast majority of patients with positive real-time RT-PCR results will eventually have typical radiological features. In addition, it was not difficult to discern COVID-19 because of its specific radiological abnormalities on chest CT image^[22, 23]^.

Therefore, CT should be used as the basic diagnostic method for COVID-19. A CT re-examination of patients without any clear clinical features at intervals of about 5 days would give a clear radiological sign and thus improved the positive rate of diagnosis.

The included individuals in this study were all patients with positive real-time RT-PCR results. All of them had different clinical manifestations and 98% had typical radiological findings indicating that real-time RT-PCR detection may be a diagnostic method with a low misdiagnosis rate. However, the positive rate of the first real-time RT-PCR test was only 66%. Thus, the rate of missed diagnosis is relatively high if only use single real-time RT-PCR test^[24]^. The rate of right diagnosis is 88% for CT examination at the first time and all first-time diagnosed patients had fever and cough. RNA detection combined with CT at the first diagnosis, along with clinical manifestations can greatly increase the sensitivity and reduce the rate of missed diagnosis. The cumulative positive rate of three times of throat swabs virus RNA tests was 96%, and the cumulative diagnostic coincidence rate of two times CT tests and nucleic acid tests reached 98%. The simultaneous application of two methods can basically detect all potential patients. Based on this, we inferred that “3+2” combined strategy can diagnose most SARS-CoV-2 infected patients, which had great reference value for the diagnosis of “clinically diagnosed cases”. The application of the “3+2” diagnostic method was conducive to the effective control of the source of infection and was a key point to improve SARS-CoV-2 prevention and control.

It has showed that SARS-CoV-2 is similar with SARS-CoV and MERS-CoV, using S-protein interacts with human ACE2, thereby infecting human respiratory epithelial cells, causing immune and inflammatory responses, and then producing cytokines and inflammatory mediators, such as IL1β, IFNγ, IP10, MCP1, *et al*^[25]^. Moreover, studies have shown that the levels of GCSF, IP10, MCP1, MIP1A, and TNFα are significantly higher in patients with severe symptoms (ICU patients) than patients with mild symptoms, suggesting that the “cytokine storm” may be related to the severity of the disease^[26, 27]^. Our study showed that the levels of C-reactive protein was significantly increased in COVID-19 patients (66 [68%] of 97 patients), and ESR was significantly accelerated (49 [57.6%] of 85 patients), indicating that there is a significant inflammatory response among COVID-19 patients. Blood routine results showed a decrease in lymphocytes count (56 [54.9%] of 102 patients), which means that the acute immune response consumes immune cells and suppresses cellular immune function, especially T lymphocyte function when the body encounters a new unknown virus invasion^[28]^. Our studies also revealed that most patients had elevated ferritin levels (12 [92.3%] of 13 patients), and some patients had abnormal liver function and elevated muscle enzymes, which indicated the damage of lung, liver and other organ, and enhanced tissue repairment. Previous studies did not find an increase in ferritin among COVID-19 patients. The phenomena of elevating levels of ferritin may make it a new indicator of the severity of the disease’s tissue damage and inflammation. However, whether C-reactive protein, ESR, serum ferritin elevation, and lymphocyte reduction and their abnormalities are related to the severity of the disease merits further study.

COVID-19 patients in our hospital all received antiviral and symptomatic treatment, anti-inflammatory treatment and oxygen therapy if necessary. Symptoms released markedly after one-week treatment on most patients. The rate of transferring to ICU and death was 8% and 3%, respectively, which was significantly lower than previous studies in Wuhan city. The improvement in clinical outcome revealed that an early prevention and timely treatment could effectively reduce the rate of critical illness and fatality proven in Xiangyang region with relatively sufficient medical resources.

This study is a cross-sectional study, which only described and analyzed COVID-19 patients with positive real-time RT-PCR results. However, concerning the low sensitivity of throat swabs virus RNA detection in previous studies^[20, 21, 24]^, we speculated that a part of suspected patients or undiagnosed patients might be COVID-19 positive cases. Thus, studies excluding those patients may not actually reflect the real picture of clinical and epidemiological feature of SARS-CoV-2 infected population. Also, accounting for the limited follow-up duration, our data may not reflect the complete course of COVID-19 development. In addition, it is a cross-sectional study with limited ability to infer the causal associations. In the current state of emergency, we hoped that our data-based findings could provide help for disease prevention, control, diagnosis and treatment.

## Conclusion

Patients in Xiangyang were mostly secondary transmission cases with the family-clustering feature and unclear contact history. The rate of severe illness and death were low, whereas some patients had longer incubation period. We, therefore, recommend prolonging the quarantine period to three weeks when necessary. All patients with positive real-time RT-PCR results had significant clinical symptoms and radiological features, which suggests we could use a combination of real-time RT-PCR, chest CT scans and clinical manifestations on admission to increase the diagnostic accuracy. Sensitivity of three rounds of real-time RT-PCR together with two rounds of chest CT scans was higher compared with other diagnostic methods. Thus, “3+2” strategy should be used to increase the accuracy of diagnosis, thereby controlling the source of infection more effectively.

## Data Availability

Data are all available on the hospital's e-system

## Reference

1. Guan W, Xian J. The progress of 2019 Novel Coronavirus (2019-nCoV) event in China. J Med Virol, 2020; doi: 10.1002/jmv.25705

2. Zhu N, Zhang D, Wang W, et al. A Novel Coronavirus from Patients with Pneumonia in China, 2019. N Engl J Med, 2020; doi: 10.1056/NEJMoa2001017

3. World Health Organization. Novel Coronavirus (2019-nCoV) Situation Report-22. World Health Organization, 2020, February 11, 2020; https://www.who.int/docs/default-source/coronaviruse/situation-reports/20200211-sitrep-22-ncov.pdf?sfvrsn=fb6d49b1_2

4. Paraskevis D, Kostaki EG, Magiorkinis G, et al. Full-genome evolutionary analysis of the novel corona virus (2019-nCoV) rejects the hypothesis of emergence as a result of a recent recombination event. Infect Genet Evol, 2020, 79: 104212.

5. Cowling BJ, Leung GM. Epidemiological research priorities for public health control of the ongoing global novel coronavirus (2019-nCoV) outbreak. Euro Surveill, 2020; doi: 10.2807/1560-7917.ES.2020.25.6.2000110

6. Guarner J. Three emerging coronaviruses in two decades. Am J Clin Pathol, 2020; doi: 10.1093/ajcp/aqaa029

7. Shen K, Yang Y, Wang T, et al. Diagnosis, treatment, and prevention of 2019 novel coronavirus infection in children: experts’ consensus statement. World J Pediatr, 2020; doi: 10.1007/s12519-020-00343-7

8. Jiang S, Xia S, Ying T, et al. A novel coronavirus (2019-nCoV) causing pneumonia-associated respiratory syndrome. Cell Mol Immunol,2020; doi: 10.1038/s41423-020-0372-4

9. Huang C, Wang Y, Li X, et al. Clinical features of patients infected with 2019 novel coronavirus in Wuhan, China. Lancet, 2020; doi: 10.1016/S0140-6736(20)30183-5

10. Chen N, Zhou M, Dong X, et al. Epidemiological and clinical characteristics of 99 cases of 2019 novel coronavirus pneumonia in Wuhan, China: a descriptive study. Lancet, 2020; doi:10.1016/S0140-6736(20)30211-7

11. National Health Commission Of the People’s Republic of China. Diagnosis and treatment of novel coronavirus-infected pneumonia. National Health Commission Of the People’s Republic of China, 2020; http://www.zhrmghggjwjw.com/

12. Jin YH, Cai L, Cheng ZS, et al. A rapid advice guideline for the diagnosis and treatment of 2019 novel coronavirus (2019-nCoV) infected pneumonia (standard version). Mil Med Res, 2020, 7(1): 4.

13. Riou J, Althaus CL. Pattern of early human-to-human transmission of Wuhan 2019 novel coronavirus (2019- nCoV), December 2019 to January 2020. Euro Surveill, 2020, 25(4).

14. Wang FS, Zhang C. What to do next to control the 2019-nCoV epidemic? Lancet, 2020, 395(10222): 391–393.

15. Wilder-Smith A, Freedman D O. Isolation, quarantine, social distancing and community containment: pivotal role for old-style public health measures in the novel coronavirus (2019-nCoV) outbreak. J Travel Med, 2020; doi: 10.1093/jtm/taaa020

16. Kui L, Fang YY, Deng Y, et al. Clinical characteristics of novel coronavirus cases in tertiary hospitals in Hubei Province. Chin Med J (Engl),2020; doi: 10.1097/CM9.0000000000000744

17. Lu H. Drug treatment options for the 2019-new coronavirus (2019-nCoV). Biosci Trends, 2020; doi: 10.5582/bst.2020.01020

18. Stein RA. The 2019 Coronavirus: Learning Curves, Lessons, and the Weakest Link. Int J Clin Pract, 2020: e13488.

19. Wang J, Zhao S, Liu M, et al. ACE2 expression by colonic epithelial cells is associated with viral infection, immunity and energy metabolism. MedRexiv, 2020; doi: https://doi.org/10.1101/2020.02.05.20020545.

20. Lin C, Ye R, Xia YL. A meta-analysis to evaluate the effectiveness of real-time PCR for diagnosing novel coronavirus infections. Genet Mol Res, 2015, 14(4): 15634–41.

21. Poon LL, Chan KH, Wong O K, et al. Early diagnosis of SARS coronavirus infection by real time RT-PCR. J Clin Virol, 2003, 28(3): 233–288.

22. Kanne JP. Chest CT Findings in 2019 Novel Coronavirus (2019-nCoV) Infections from Wuhan, China: Key Points for the Radiologist. Radiology, 2020; doi: 200241. 10.1148/radiol.2020200241

23. Lei J, Li J, Li X, et al. CT Imaging of the 2019 Novel Coronavirus (2019-nCoV) Pneumonia. Radiology, 2020:200236; doi: 10.1148/radiol.2020200236

24. Corman VM, Landt O, Kaiser M, et al. Detection of 2019 novel coronavirus (2019-nCoV) by real-time RT-PCR. Euro Surveill, 2020, 25(3).

25. Wu A, Peng Y, Huang B, et al. Genome Composition and Divergence of the Novel Coronavirus (2019-nCoV) Originating in China. Cell Host Microbe, 2020; doi: 10.1016/j.chom.2020.02.001

26. Chu D, Pan Y, Cheng S, et al. Molecular Diagnosis of a Novel Coronavirus (2019-nCoV) Causing an Outbreak of Pneumonia. Clin Chem, 2020; doi: 10.1093/clinchem/hvaa029

27. Wang Z, Chen X, Lu Y, et al. Clinical characteristics and therapeutic procedure for four cases with 2019 novel coronavirus pneumonia receiving combined Chinese and Western medicine treatment. Biosci Trends, 2020; doi: 10.5582/bst.2020.01030

28. Baruah V, Bose S. Immunoinformatics-aided identification of T cell and B cell epitopes in the surface glycoprotein of 2019-nCoV. J Med Virol, 2020; doi: 10.1002/jmv.25698

